# Within-person temporal alignment shows symptom co-fluctuations and early precursors of suicidal ideation

**DOI:** 10.64898/2026.01.27.26344922

**Authors:** A.J.C. van der Slot, C. Boonmann, M. Eikelenboom, M.W.M. Gijzen, A.A.L. Kok, D. de Beurs, B.W.J.H. Penninx, E.J. Giltay

**Author notes:** **Corresponding authors:** Abe J.C. van der Slot, Department of Psychiatry, Leiden University Medical Center, Postal box 9600, Postal zone B1-P, 2300 RC Leiden, the Netherlands, Erik J. Giltay, Department of Psychiatry, Leiden University Medical Center, Postal box 9600, Postal zone B1-P, 2300 RC Leiden, the Netherlands. Electronic address. **Source of Funding:** COVID online data collection and analyses were partly funded by a ‘fast track grant’ from the Dutch Research Council (grant no 440.20.009) and by the RESPOND project which has received funding from the European Union’s Horizon 2020 research and innovation programme Societal Challenges under grant agreement No 101016127. The infrastructure for the NESDA study is funded through the Geestkracht programme of the Netherlands Organisation for Health Research and Development (grant no 10-000-1002) and financial contributions by participating universities and mental health-care organisations (VU University Medical Center, Geestelijke Gezondheidszorg (GGZ) inGeest, Leiden University Medical Center, Leiden University, GGZ Rivierduinen, University Medical Center Groningen, University of Groningen, Lentis, GGZ Friesland, GGZ Drenthe, Rob Giel Onderzoekscentrum). The infrastructure for the NESDO study is funded through the Fonds NutsOhra (project 0701-065), Stichting tot Steun VCVGZ, NARSAD The Brain and Behaviour Research Fund (grant id 41080), and by participating universities and mental health-care organisations (VU University Medical Center, Leiden University Medical Center, University Medical Center Groningen, University Medical Center St Radboud, GGZ inGeest, GGNet, GGZ Nijmegen, GGZ Rivierduinen, Lentis, and Parnassia). The infrastructure for the NOCDA study is funded by participating universities and mental health-care organisations (Academic Department VU Medical Center, GGZ inGeest, Innova Research Centre, Mental Health Care Institute GGZ Centraal, Marina de Wolf Anxiety Research Centre, Center for Anxiety Disorders Overwaal, Dimence, GGZ Overijssel, Department of Psychiatry at Leiden University Medical Center, Vincent van Gogh Institute Mental Health Care Centre, Academic Anxiety Center, PsyQ Maastricht University, Division Mental Health and Neuroscience, and Stichting tot Steun VCVGZ). **Role of the funder:** The funders had no role in the design and conduct of the study; collection, management, analysis, and interpretation of the data; preparation, review, or approval of the manuscript; and decision to submit the manuscript for publication. **Data Availability Statement:** The data used in this study were obtained from the NESDA, NESDO, and NOCDA cohorts. Due to privacy restrictions and ethical regulations, the raw data are not publicly available. Access to the data can be requested through the respective study secretariats, subject to approval by their scientific committees and in accordance with their data access protocols. **Declaration of generative AI and AI-assisted technologies in the manuscript preparation process:** During the preparation of this work the authors used *ChatGPT 5.1* for text processing only. After using this tool, the authors reviewed, revised, and edited the content as needed and take full responsibility for the content of the published article.

## Abstract

**Background:** Suicidal ideation (SI) is a major global concern, yet its dynamic interplay with other symptoms remains poorly understood.

**Objective:** To identify symptoms that co-fluctuate with or temporally precede SI to improve warning signal detection and intervention.

**Methods:** Longitudinal data from three Dutch psychiatric cohorts with lifetime internalizing disorders (16 waves from April 2020 until February 2022) were collected during the COVID-19 pandemic. We analyzed depressive, happiness, anxiety, loneliness, worry symptoms, and COVID-19-specific items only in those participants with SI fluctuations. Dynamic Time Warping (DTW) quantified within-person similarity between symptom trajectories and SI, and results were aggregated at group level.

**Findings:** The 307 participants (mean age 44.8 years; 61.6% female) showed increasing SI over time (p < .001). SI aligned with four depressive symptoms (i.e., sad mood, low self-esteem, low interest, and reduced happiness), two anxiety-related symptoms (i.e., fear of losing control, faintness), feeling abandoned, and overwhelming worrying. In directed DTW analysis, sad mood, hypersomnia, worrying about projects, and numbness/tingling showed significant temporal precedence before SI.

**Conclusion:** SI is embedded in a broad symptom network beyond depression. These results underscore the value of time-sensitive, idiographic monitoring using tools like DTW to capture the person-specific temporal pathways through which SI emerges and intensifies.

**Clinical implications:** This study suggests a core group of affective, cognitive, and interpersonal symptoms that could serve as informative signals for evaluating changes in SI and may represent actionable targets for intervention.

**Summary Box:** *What is already known on this topic?:*

- Suicidal ideation (SI) is a dynamic phenomenon, yet traditional research often relies on static, group-level averages that do not capture individual fluctuations.
- While SI is linked to depression, it can emerge independently through complex interactions with other affective and interpersonal states

*What this study adds?:*

- This study identifies a set of affective, cognitive, and interpersonal symptoms, sad mood, overwhelming worry, and feelings of abandonment, that significantly co-fluctuate with SI over weeks and months. Additionally four specific “leading” symptoms, sad mood, hypersomnia, worrying about projects, and somatic numbness, were found that precede increases in SI.

*How this study might affect research, practice or policy?:*

- The identified co-fluctuations and precursors serve as informative “(early) warning signals” that can improve individual risk stratification and clinical monitoring and may represent targets for intervention.
- The results support a shift toward network-based models in suicidology, emphasizing the need for time-sensitive monitoring to capture the complex and dynamic nature of suicidality.

## 1. introduction

Suicidal ideation (SI) is a pressing global concern, with lifetime prevalence reaching 22.7% [1]. Although linked to depression, the disorder’s heterogeneity limits its predictive power [2], underscoring the need to examine symptom-level dynamics rather than broad diagnostic categories [3]. Evidence shows SI can arise independently of depressive severity, suggesting mechanisms beyond depression alone must be studied [4]. Other affective and interpersonal states, such as anxiety and loneliness, are also recognized as proximal risk factors [5]. Theoretical models, like the integrated motivational-volitional (IMV) model [6], support this by conceptualizing SI as emerging from dynamic interactions between factors like entrapment, fluctuating affect and interpersonal stress [7]. However, it remains largely unclear how SI fluctuates over time and which symptoms co-fluctuate with its course, limiting timely detection and intervention [8]. The COVID-19 pandemic further highlighted this; while it intensified risk factors like distress and loneliness [9] SI trends were inconsistent. Some studies reported rising SI [10], while others found it remained stable despite rising depression and anxiety symptoms [11]. These divergent patterns indicate that SI does not simply track general distress, highlighting the need for approaches that capture its dynamic interplay with co-occurring symptoms and stressors. Additionally, such dynamics may differ across gender and age groups. Gender-specific patterns in risk expression suggest that different symptoms may drive the course of SI [12]. In a cross sectional network study in younger women, SI was closely linked to hopelessness and loneliness [13], whereas in men, low self-worth and perceived loss of control were more central [14].

In this study, we applied Dynamic Time Warping (DTW), a nonlinear alignment algorithm that detects temporal patterns in intra-individual symptom trajectories despite irregular sampling and varying sequence lengths [15]. DTW is particularly suited for mental health research, where the assumption that group-level patterns mirror individual processes (ergodicity) is frequently violated [16, 17]. DTW can also assess temporal directionality, providing preliminary insight into which symptoms may precede or follow changes in SI [15]. Recent work in psychopathology has emphasized that mental disorders emerge from dynamic symptom interactions rather than static traits [17, 18]. Network models provide a useful framework by conceptualizing symptoms as interconnected nodes with potential temporal directionality [18]. Early psychiatric applications of DTW have demonstrated its value for modelling personalized dynamics, yet only two studies have examined SI at the individual level [7, 19]. Building on ecological momentary assessment (EMA) research showing that SI and its proximal risk factors fluctuate markedly within individuals over hours to days [20, 21], these studies applied DTW to EMA data, identifying key roles for entrapment and rumination and showing that low inner peace preceded suicidality. However, their small sample sizes (n = 11 and n = 28) limited generalizability beyond short-term within-day fluctuations. Therefore, in this study we investigate which affective, cognitive and interpersonal symptoms co-fluctuate with SI across weeks and months, whether certain symptoms temporally precede changes in SI, and whether these alignments differ by sex and age. Mapping these dynamics can deepen our understanding of the mechanisms underlying suicidality and may provide novel approaches for uncovering the dynamic processes through which suicidality emerges and intensifies.

## 2. methods

### 2.1 Study Population and Procedure

Data were integrated from three Dutch longitudinal psychiatric cohorts: the Netherlands Study of Depression and Anxiety (NESDA; n = 2,329) [22], the Netherlands Study of Depression in Older Persons (NESDO; n = 510) [23], and the Netherlands Obsessive Compulsive Disorder Association Study (NOCDA; n = 419) [24]. These cohorts employ harmonized diagnostic procedures and data collection protocols; detailed descriptions of the study designs, recruitment procedures, and assessment batteries are provided elsewhere [22-24].

### 2.2 Participants

During the COVID-19 pandemic (April 2020–February 2022), 2,748 participants were invited for up to sixteen waves of online questionnaires. Of the 1,125 individuals who completed at least one assessment, we included those with:

1. Complete data for at least four waves on all relevant items.
2. At least one within-person fluctuation in suicidal ideation (SI; ≥ point change on QIDS-SR16 item 12).

The final analytical sample (n=307) consisted of participants from NESDA (n=252), NESDO (n=11), and NOCDA (n=44).

While included and excluded participants did not differ in age, sex, or educational level, the included sample showed significantly higher comorbidity (see Supplementary Table 1). Supplementary Figure 1 shows the inclusion/exclusion flowchart. The study protocol was approved by the Institutional Review Board of the Vrije Universiteit Medical Center, Amsterdam (ref. 2020.166), and participants provided informed consent digitally.

### 2.3 Instruments

Mental health outcomes during both pre-pandemic and COVID-19 measurement waves included:

Suicidal ideation (SI) was assessed with item 12 of the 16-item QIDS-SR_16_. Response options range from 0 = ‘I do not think of suicide or death’, 1 = ‘I feel that life is empty or wonder if it is worth living’, 2 = ‘I think of suicide or death several times a week for several minutes’, to 3 = ‘I think of suicide or death several times a day in depth, or have made specific plans, or have actually attempted suicide’. This item served as the primary dependent variable.

Depressive symptoms were assessed with the remaining 15 items of the QIDS-SR_16_, (α ≈ 0.81–0.86 [25].

Anxiety symptoms were assessed with the 21-item Beck Anxiety Inventory (BAI; α ≈ 0.92– 0.94) [26].

Loneliness symptoms were measured with the six-item De Jong Gierveld Loneliness Scale (DJGLS; α ≈ 0.77–0.79) [27].

Worry was assessed using the 11-item Penn State Worry Questionnaire (PSWQ-11; α ≈ 0.93) [28].

Happiness was evaluated with the single-item Self-Rating of Happiness scale (SRH; range 1– 7), reverse-coded for analysis so higher scores indicate lower happiness [29].

COVID-19 Stressors were captured via 21 items covering emotional impact, coping, and adherence to measures (on a 5-point Likert scale)[30].

### 2.4 Covariates

Baseline sociodemographic covariates included age, sex, and years of education using in the mixed models to yield the two forest plots. Clinical variables comprised lifetime diagnoses of major psychiatric disorders (mayor depressive disorder (MDD), general anxiety disorder (GAD), panic disorder, and obsessive compulsive disorder (OCD)) established via the Composite International Diagnostic Interview (CIDI) or Structured Clinical Interview for DSM-IV (SCID-I)[22-24].

Two indices of psychiatric burden were computed: (1) the total number of lifetime disorders, reflecting diagnostic comorbidity, and (2) chronicity, categorized as no current disorder, ≤50% of preceding waves with a current disorder, or >50% of waves with a current disorder.

### 2.5 statistical analysis

Baseline characteristics were summarized using means (standard deviation (SD)), medians (inter quartile range (IQR)), or counts (%) as appropriate, stratified by age and sex. Temporal trends in SI over the 22-month pandemic period were visualized alongside national COVID-19 mortality data and tested using a linear mixed-effects model with time as a fixed effect and participant as a random intercept

Prior to analysis, all items were standardized (z-scores) and harmonized so that higher scores consistently represented greater severity. To ensure robustness against outliers and irregular sampling, scores were winsorized (capped at ±3). To optimize the DTW alignment, five intermediate points were linearly interpolated between each pair of consecutive assessments. This step was taken specifically to increase the temporal resolution of the trajectories, thereby reducing artifacts caused by mismatched start and endpoints, rather than to impute missing primary data.

To examine the temporal alignment between SI and other variables, we first applied an undirected DTW to participant-level panel data. DTW is a non-linear alignment technique that quantifies the similarity between two time-series by identifying the optimal match between their trajectories, even when the timing of fluctuations differs [31]. This approach captures parallel within-person changes regardless of irregular spacing between assessments. A time-window constraint (Sakoe–Chiba band) of 1 allowed each observation to align with points one wave earlier or later, emphasizing co-fluctuation occurring at or near the same assessment [15]. The method is well suited for the relatively short and unevenly spaced series collected here (16 waves per participant over 22 months). For each participant, pairwise DTW distances were computed across all 74 items. This yielded 307 individual (74 by 74) DTW distance matrices. Throughout, we use the term ‘symptoms’ to refer to all included items, acknowledging that the COVID-19 variables represent contextual stressors and behaviours. At the group level, each symptom pair’s DTW distance was compared against the average distance of all other symptom pairs. P-values tested whether a pair’s distance was significantly smaller than the empirical mean, corresponding to the null hypothesis of independent fluctuation. Only symptom pairs with significantly smaller distances (p < .05) were retained. This undirected network reflects unadjusted associations and served as the descriptive basis for the adjusted mixed-effects analyses presented in the forest plots and supplementary figures.

Linear mixed-effects models, adjusting for age, sex, and education, estimated the relative strength of alignment between SI and each symptom. Random intercepts accounted for participant-level clustering. Although our hypotheses concerned only smaller distances to SI (i.e., a one-sided alternative), we applied two-sided testing to account for the large number of statistical tests across all 73 symptoms and to adopt a conservative approach.

To further explore temporality, we applied a directed DTW analysis using an asymmetric Sakoe–Chiba window to estimate lead–lag relationships (Kopland & Giltay, 2025). For each participant, directed DTW matrices were computed and then averaged per symptom to obtain group-level directionality estimates. One-sample *t*-tests against zero (two-tailed, *p* < .05) identified symptoms that significantly preceded or followed changes in SI. Out-strength (temporal lead) and in-strength (temporal lag) indices summarized these effects.

All statistical analyses were performed using the packages “dtw” (version 1.231), “parallelDist” (version 0.2.6), “forestplot” (version 3.1.3) and “qgraph” (version 1.9.8) of the statistical program R Studio version 4.3.2; R Foundation for Statistical Computing, Vienna, Austria, 2016. URL:https://www.R-project.org/

## 3. Results

### 3.1 Patient characteristics

The 307 included participants completed an average of 10.7 assessments (SD = 3.71, range = 4–16) and had a mean age of 44.8 years (SD = 11.2), with 61.6% being female and 49.5% having completed a high level of education. Lifetime diagnoses were common in this group, with all having a history of a mental disorder, and high rates of lifetime MDD (65.8%), GAD (31.4%), and panic disorder (32.7%). Regarding functional outcomes during the pandemic, the sample had significantly higher scores on average depression (QIDS), anxiety (BAI), and loneliness (PWSQ) scores, compared to excluded participants (all p < .001). Included participants also had more lifetime disorders and had a higher chronicity of disorders compared to excluded individuals (both p < .001), as assessed through CIDI/SCID-based diagnostic interviews. Table 1 shows the sociodemographic and clinical characteristics of the 4 demographic subgroups defined by age and sex (n=307).

**Table 1.**
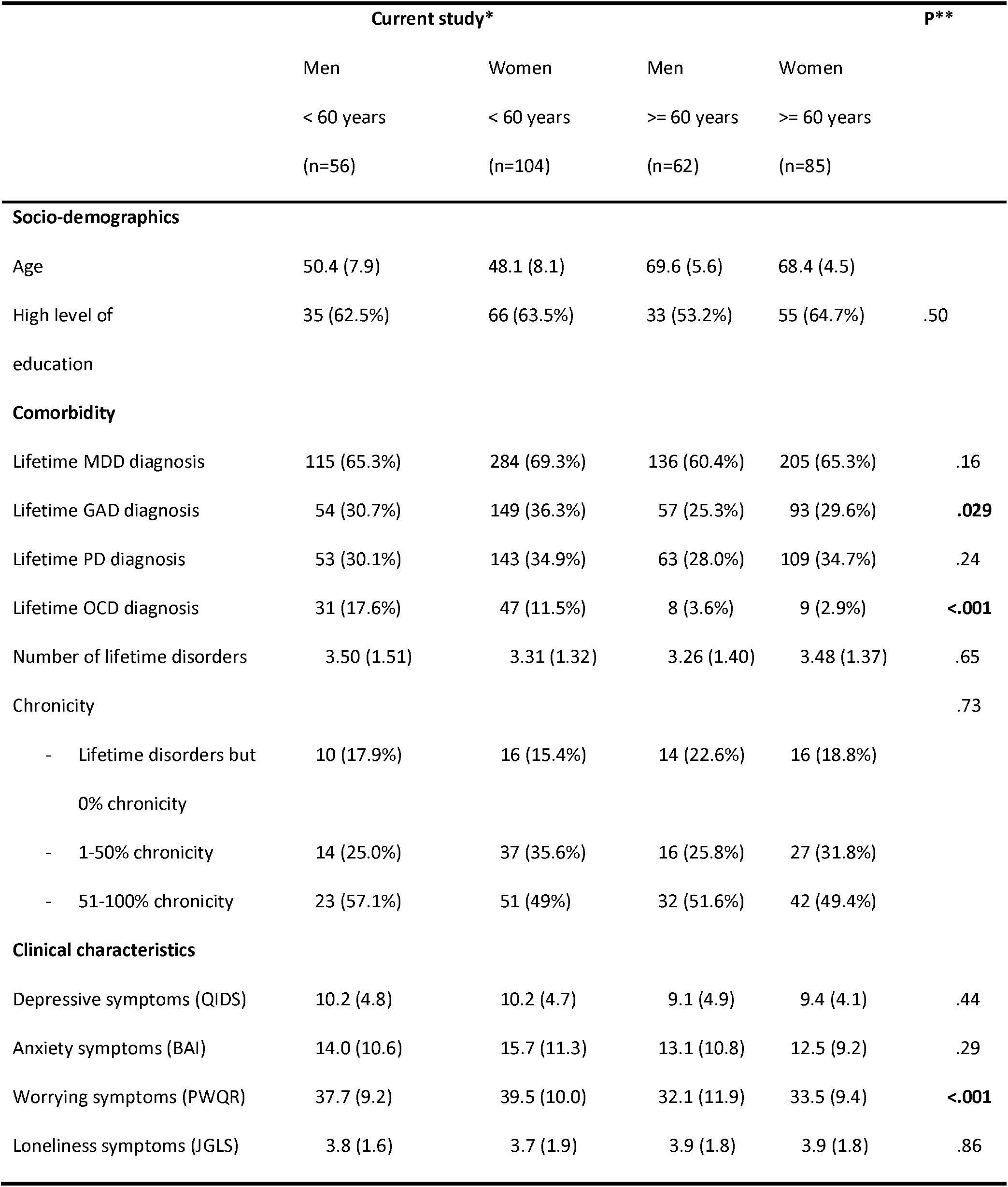
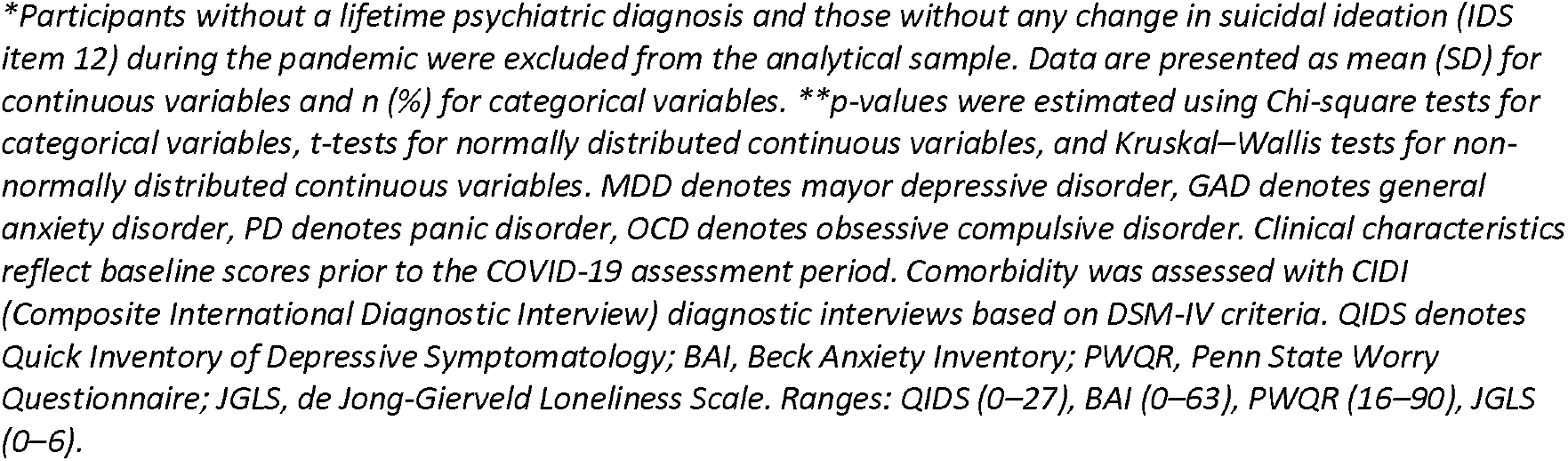
Comparison of baseline characteristics in the four demographic subgroups of the sample (307 participants)

|  | Current study* |  |  |  | p** |
| --- | --- | --- | --- | --- | --- |
|  | Men | Women | Men | Women |  |
|  | < 60 years | < 60 years | >= 60 years | >= 60 years |  |
|  | (n=56) | (n=104) | (n=62) | (n=85) |  |
| **Socio-demographics** |  |  |  |  |  |
| Age | 50.4 (7.9) | 48.1 (8.1) | 69.6 (5.6) | 68.4 (4.5) |  |
| High level of education | 35 (62.5%) | 66 (63.5%) | 33 (53.2%) | 55 (64.7%) | .50 |
| **Comorbidity** |  |  |  |  |  |
| Lifetime MDD diagnosis | 115 (65.3%) | 284 (69.3%) | 136 (60.4%) | 205 (65.3%) | .16 |
| Lifetime GAD diagnosis | 54 (30.7%) | 149 (36.3%) | 57 (25.3%) | 93 (29.6%) | **.029** |
| Lifetime PD diagnosis | 53 (30.1%) | 143 (34.9%) | 63 (28.0%) | 109 (34.7%) | .24 |
| Lifetime OCD diagnosis | 31 (17.6%) | 47 (11.5%) | 8 (3.6%) | 9 (2.9%) | **<.001** |
| Number of lifetime disorders | 3.50 (1.51) | 3.31 (1.32) | 3.26 (1.40) | 3.48 (1.37) | .65 |
| Chronicity |  |  |  |  | .73 |
| - Lifetime disorders but 0% chronicity | 10 (17.9%) | 16 (15.4%) | 14 (22.6%) | 16 (18.8%) |  |
| - 1-50% chronicity | 14 (25.0%) | 37 (35.6%) | 16 (25.8%) | 27 (31.8%) |  |
| - 51-100% chronicity | 23 (57.1%) | 51 (49%) | 32 (51.6%) | 42 (49.4%) |  |
| **Clinical characteristics** |  |  |  |  |  |
| Depressive symptoms (QIDS) | 10.2 (4.8) | 10.2 (4.7) | 9.1 (4.9) | 9.4 (4.1) | .44 |
| Anxiety symptoms (BAI) | 14.0 (10.6) | 15.7 (11.3) | 13.1 (10.8) | 12.5 (9.2) | .29 |
| Worrying symptoms (PWQR) | 37.7 (9.2) | 39.5 (10.0) | 32.1 (11.9) | 33.5 (9.4) | **<.001** |
| Loneliness symptoms (JGLS) | 3.8 (1.6) | 3.7 (1.9) | 3.9 (1.8) | 3.9 (1.8) | .86 |

*\*Participants without a lifetime psychiatric diagnosis and those without any change in suicidal ideation (IDS item 12) during the pandemic were excluded from the analytical sample. Data are presented as mean (SD) for continuous variables and n (%) for categorical variables. \*\*p-values were estimated using Chi-square tests for categorical variables, t-tests for normally distributed continuous variables, and Kruskal–Wallis tests for non-normally distributed continuous variables. MDD denotes major depressive disorder, GAD denotes general anxiety disorder, PD denotes panic disorder, OCD denotes obsessive compulsive disorder. Clinical characteristics reflect baseline scores prior to the COVID-19 assessment period. Comorbidity was assessed with CIDI (Composite International Diagnostic Interview) diagnostic interviews based on DSM-IV criteria. QIDS denotes Quick Inventory of Depressive Symptomatology; BAI, Beck Anxiety Inventory; PWQR, Penn State Worry Questionnaire; JGLS, de Jong-Gierveld Loneliness Scale. Ranges: QIDS (0–27), BAI (0–63), PWQR (16–90), JGLS (0–6).*

### 3.2 Sample trends in SI

Figure 1 displays temporal trends in SI across the sample (n = 307). Mean SI scores increased significantly over time (p < 0.001, top panel), while the bottom panel shows weekly COVID-19 mortality in the Netherlands annotated with major public health measures (e.g., lockdowns, curfews). Only a slight correspondence was observed between temporary increases in mean SI and peaks in COVID-19 deaths or restrictions.

**Fig. 1.**
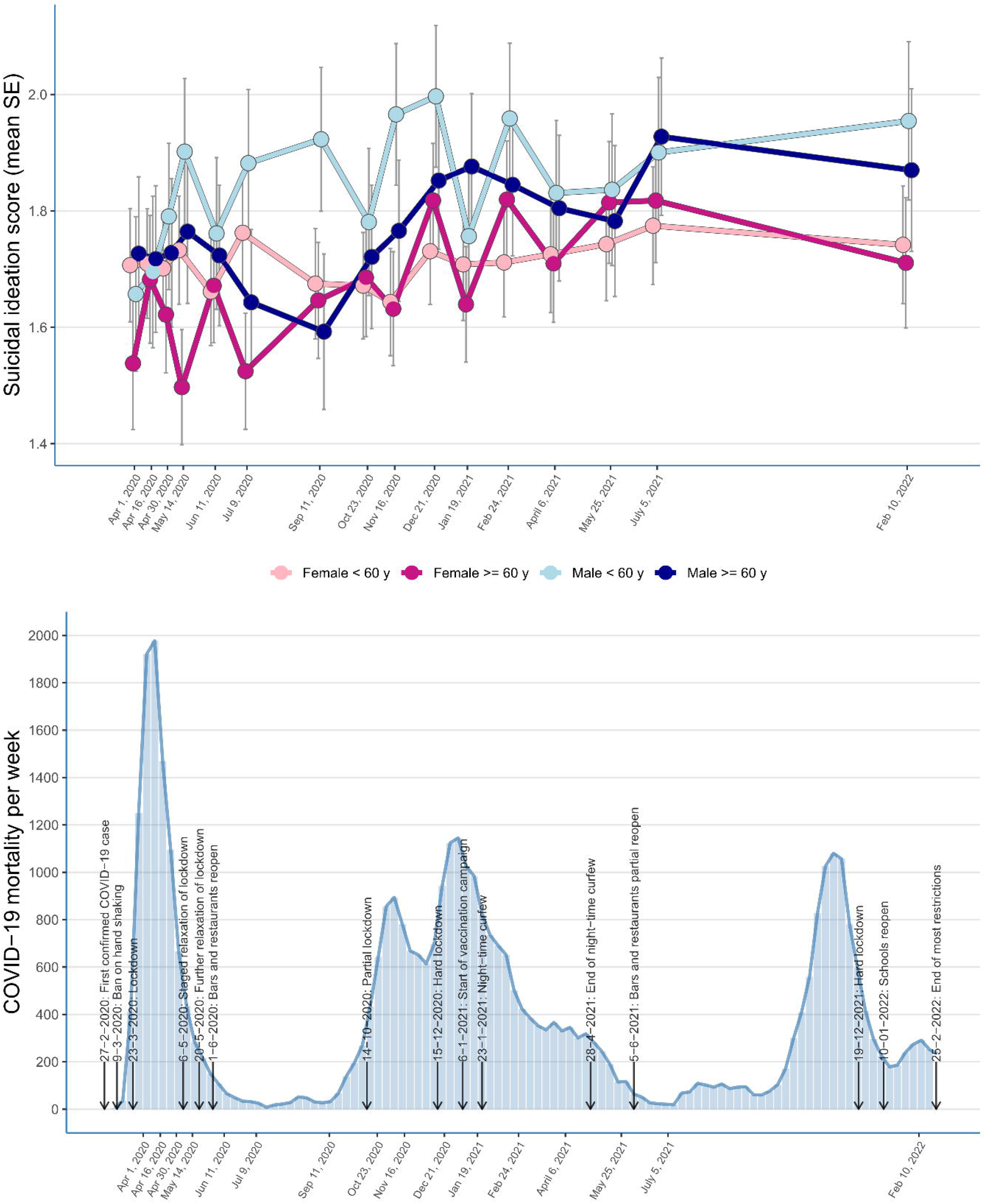

### 3.3 Undirected symptom network structure

Figure 2 presents the undirected DTW symptom network, retaining only edges between symptom pairs with significantly smaller DTW distances than expected. The network shows moderate clustering of symptoms within their original scales (e.g., IDS, BAI, PSWQ), but also notable cross-domain connections, suggesting interrelations across symptom domains. Suicidal ideation (IDS12), highlighted in red, was positioned at the periphery of the network, and maintained connections with key symptoms from multiple domains. Significant connections were observed with depressive symptoms, including low interest (IDS13), low self-esteem (IDS16), and sad mood (IDS5), as well as with the loneliness symptoms “feeling abandoned” (L6) and “feeling lonely” (L5). Additional connections were found with cognitive-affective anxiety symptoms “fear of losing control” (BAI14), “faintness” (BAI19) and reduced happiness. In contrast, COVID-specific symptoms clustered peripherally, with no direct connections to SI.

**Fig. 2.**
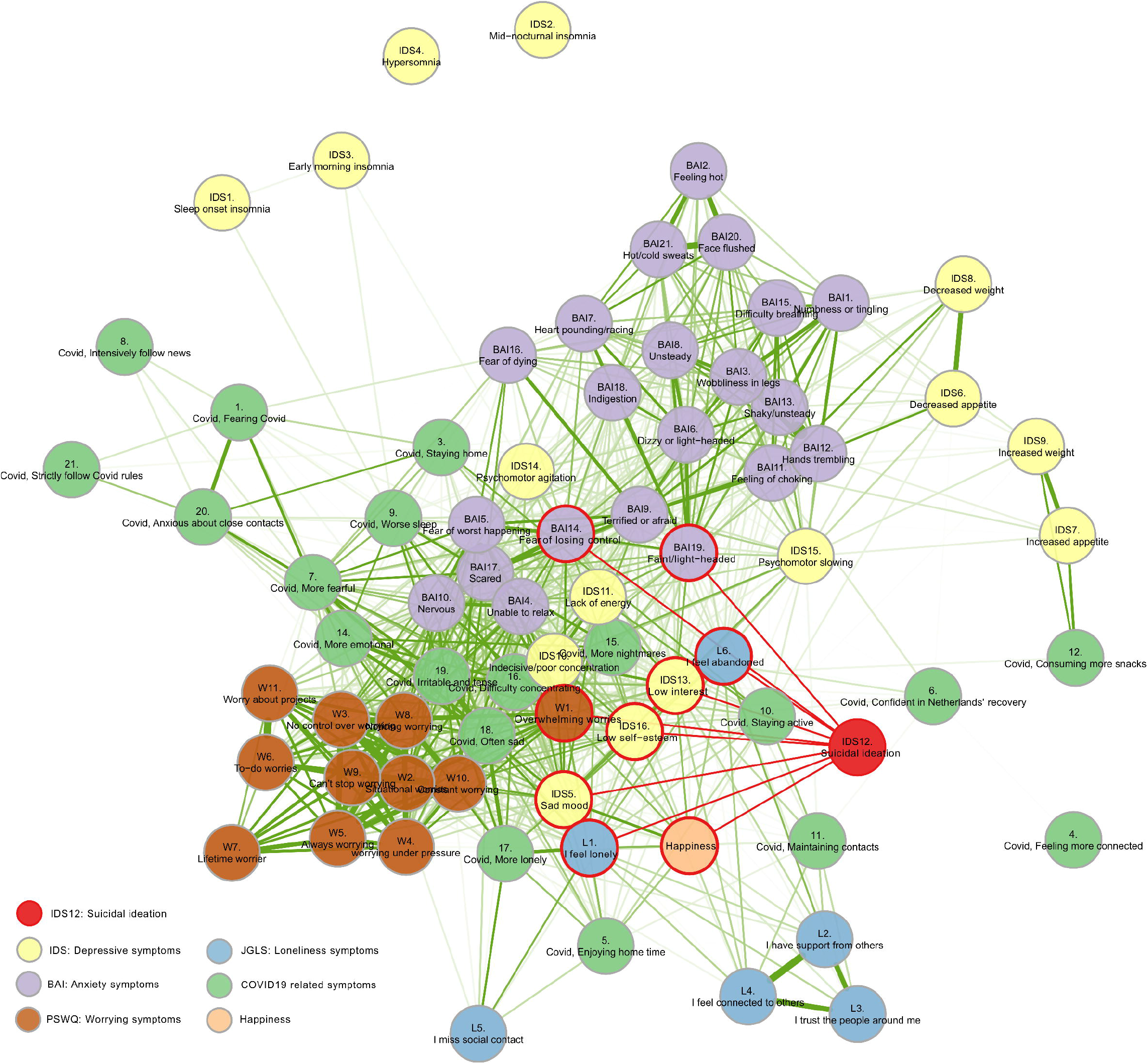

### 3.4 Symptom-specific alignments with SI (forest plot results)

We examined DTW distances between SI and each of the 73 symptoms, adjusting for age, sex, and educational level, to identify which symptoms co-fluctuated most closely with SI. Results are displayed in a forest plot (see Figure 3), showing the relative strength of alignment between SI and individual symptoms. Smaller DTW distances indicate stronger temporal alignment with SI, and thus indicate symptoms that co-vary more closely with SI. Several affective symptoms (i.e. “sad mood”, “low self-esteem”, and “low interest”) together with the separate happiness symptom, showed significant alignment with SI. Two anxiety symptoms (“fear of losing control” and “feeling faint or light-headed”) and the cognitive symptom “overwhelming worrying” were also significantly aligned with SI. In addition, the loneliness symptom “I feel abandoned” co-fluctuated significantly with SI. By contrast, none of the COVID-specific and somatic symptom showed significant alignment.

**Fig. 3.**
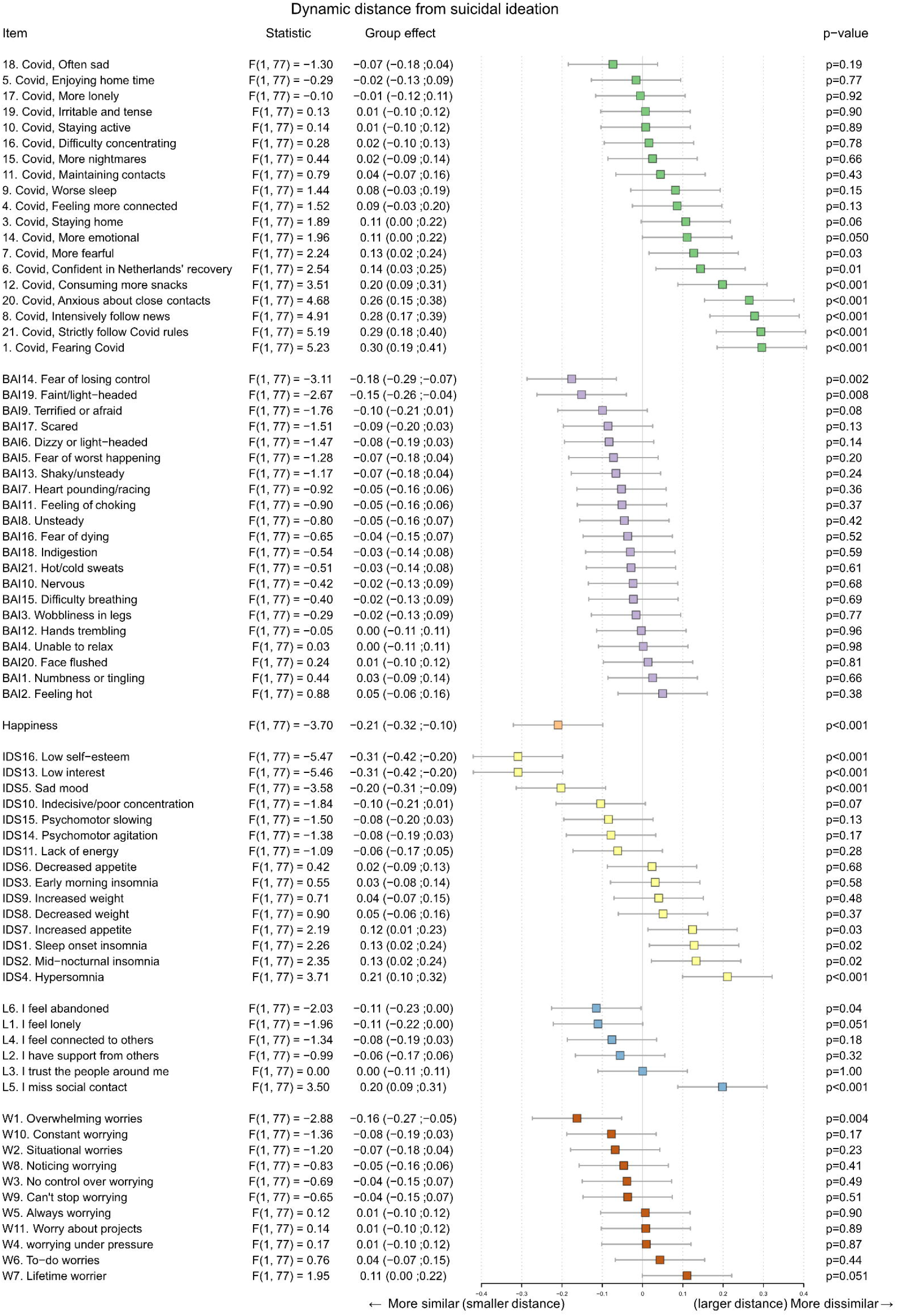
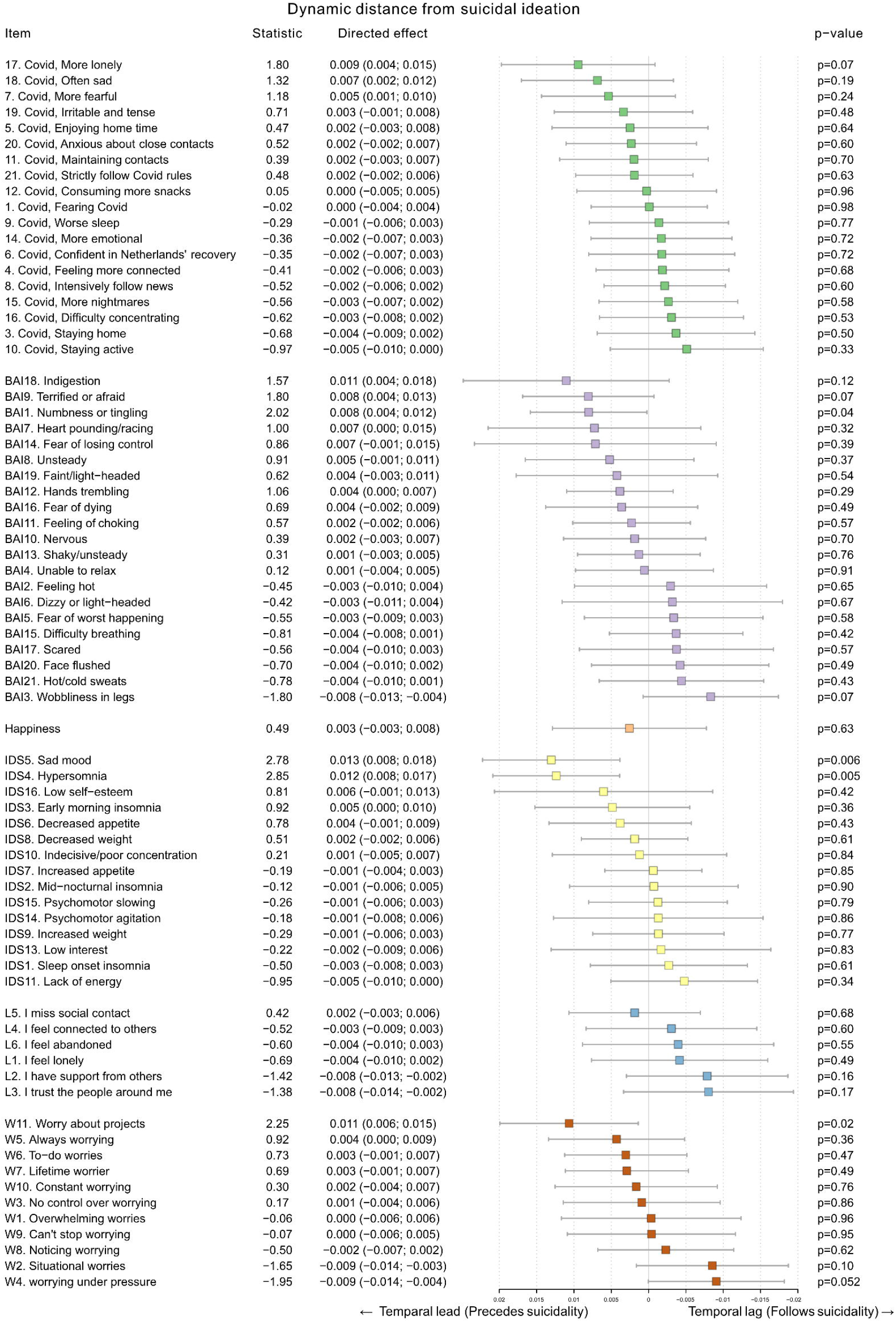

Post-hoc subgroup analyses by sex and age indicated no significant effect modification of these associations, suggesting consistency across demographic groups (Supplementary Figures 2–6 for detailed results).

#### 3.5 Directed temporal associations with SI

Figure 3 shows the forest plot displaying the directed DTW results, indicating which symptoms tended to statistically significantly precede or follow fluctuations in suicidal ideation (SI) at the group level. Positive directed values reflect temporal lead, symptoms that typically rise before increases in SI, whereas negative values reflect temporal lags.

Four symptoms showed significant leading associations with SI: sad mood, hypersomnia, worrying about projects, and BAI1 numbness/tingling, indicating that increases in these symptoms tended to precede subsequent increases in SI. No symptoms demonstrated significant lagging effects, suggesting that no specific symptom reliably followed changes in SI.

## 4. Discussion

The aim of the current study was to map the dynamic, within-person interplay between suicidal ideation and a broad range of affective, cognitive, and interpersonal symptoms. The study identified a consistent pattern of symptom co-fluctuation with SI, clustering around affective dysregulation (i.e. sad mood, low self-esteem, low interest, reduced happiness), cognitive overload (i.e. overwhelming worry, fear of losing control, faintness), and interpersonal distress (i.e. feelings of abandonment). None of the COVID-19-specific stressors and concerns showed a significant temporal alignment with SI, underscoring the greater role of internal psychological processes. In the directed analysis, sad mood, hypersomnia, worrying about projects and somatic numbness significantly preceded subsequent changes in SI.

The alignment of SI with core affective symptoms such as sadness, low self-worth, and low interest supports the view of suicidality as a fluctuating, state-like phenomenon shaped by proximal emotional experiences [3, 20]. The alignment with overwhelming worry, fear of losing control, and faintness suggests that acute experiences of emotional and cognitive overwhelm or loss of agency could trigger SI [32], consistent with implicit cognition studies suggesting that some individuals perceive possible death as a means of regaining control [33]. Our most notable finding was the specific alignment of SI with the feeling of being abandoned, which reflects perceptions of interpersonal rupture rather than generalized loneliness. Interpersonal frameworks likewise emphasize that thwarted belongingness and perceived burdensomeness heighten SI [34], and reviews identify rejection, defeat, entrapment, and humiliation as particularly salient social precipitants of suicidality [35]. While loneliness and social isolation have been shown to predict SI [36], our findings suggest that feeling abandoned was more proximally linked to suicidal ideation, consistent with later-life research on the relationship between perceived loss of social worth and abandonment on SI [37]. Although pandemic-related stressors have been linked to SI elsewhere [9], our analyses showed no temporal alignment of the COVID-19 symptoms with SI. This suggests their effects may be small, more distal or could be mediated via symptoms more proximal to SI like sadness, panic or feelings of abandonment [10]. Together, these results emphasize that SI arises from dynamic interplay between affective, cognitive, and interpersonal states, whereas external stressors appear to act indirectly through these emotional processes.

Beyond the undirected results, the directed DTW analysis identified a subset of symptoms showing temporal precedence over SI. Although effect sizes were small, the leading role of mood and sleep aligns with evidence that these changes act as early indicators of rising suicidality [20, 38]. Similarly, the precedence of worry and somatic symptoms (numbness/tingling) is consistent with their roles as robust cognitive and autonomic risk factors [39, 40]. The lack of more consistent leading effects across the entire symptom set likely reflects the high heterogeneity of suicidal processes, where suicidality emerges through multiple person-specific temporal pathways [8].

### Implications

This study demonstrates the utility of DTW for psychiatric research, as it accommodates irregular, non-stationary data typical of naturalistic cohorts [31]. Our findings show that even with lower-frequency assessments, DTW can recover meaningful symptom co-fluctuations, complementing EMA studies that capture within-hours/days SI dynamics [20]. Prior DTW-based EMA studies in very small samples have shown that entrapment, worry, sadness, low self-worth, loneliness, and low inner peace often precede short-term increases in SI [7, 19], consistent with the Integrated Motivational-Volitional model [6]. Our findings, based on a larger cohort spanning weeks/months, bridge short- and longer-term timescales and reinforce models viewing SI as a state-like process driven by proximal emotions [5]. More broadly, these results underscore the importance of idiographic, within-person approaches, as group-level associations do not necessarily generalize to individual processes [17]. They support a shift toward models that conceptualize suicidality as emerging from dynamic symptom networks rather than fixed latent constructs [8, 18]. Integrating DTW with network modelling or digital phenotyping may further enhance risk prediction and contribute to real-time early warning systems [18].

Clinically, our findings provide a fine-grained understanding of how suicidality is reinforced by dynamic symptom processes. The identified co-fluctuations suggest that affective, cognitive, and interpersonal symptoms can serve as informative signals when evaluating changes in suicide risk in high-risk populations. Recognizing and discussing these co-occurring symptoms may also help reduce stigma and facilitate conversations about suicidality. For intervention, these symptoms represent actionable targets; for instance, increases in worry, perceived loss of control, or feelings of abandonment can inform safety planning and tailored therapeutic focus [10]. Additionally, early indicators such as hypersomnia, worrying about projects, or somatic numbness may support earlier recognition of escalating risk within this psychiatric context. Digital tools may facilitate such monitoring. EMA enables high-frequency tracking of affect and cognition, while passive sensing can capture abrupt behavioural shifts with minimal patient burden [41]. Tools such as the Prior Elicitation Module for Idiographic System Estimation (PREMISE) integrate clinician knowledge with individualized symptom networks to improve interpretability [42]. Whereas PREMISE builds on vector autoregression to estimate predictive relations among symptoms, DTW captures synchrony in symptom trajectories without assuming fixed temporal lags. Together, these complementary approaches illustrate how dynamic symptom monitoring may contribute to more personalized and clinically meaningful suicide risk assessment [7, 20].

### Limitations & Strengths

Several limitations should be noted. First, although the number of assessments was sufficient, their spacing was irregular, ranging from biweekly to several months. Such variability complicates time-series analysis and may have obscured very rapid, short-term fluctuations in SI that occur between waves. However, DTW is less adversely affected by non-stationary and unevenly spaced data than parametric methods such as Multilevel Vector AutoRegression (mlVAR) [31]. Second, participants were middle aged and older individuals with a West-European ethnic background and lifetime affective disorder histories, limiting generalizability to younger and other ethnic groups of subjects [43]. Third, requiring ≥4 assessments may have led to a selection bias toward more compliant participants. Fourth, symptom alignment was assessed using relative DTW distances rather than absolute proximity, and directed effects at the group level were small, likely due to substantial between-person heterogeneity and measurement error of assessing individual symptoms through single items. Lastly, while DTW captures temporal synchrony between symptoms, it does not permit causal inference [44].

Despite these limitations, the study has several strengths. First, it applied DTW to model intra-individual symptom dynamics in a large harmonized cohort, providing a naturalistic test of SI trajectories during the COVID-19 pandemic. Second, the within-person modelling approach aligns with calls for idiographic, temporally sensitive models in psychiatry [45]. Third, participants completed on average 10.7 assessments, offering a solid basis for panel data analyses using DTW.

## Conclusion

In conclusion, this study identified a consistent set of affective and cognitive symptoms that co-fluctuate with SI across weeks to months, regardless of age or sex, whereas COVID-19-specific stressors played only a limited role. Sad mood, hypersomnia, worrying about projects and numbness may precede increases in SI, but the absence of other significant leading effects indicates that temporal pathways into suicidality vary considerably across individuals. Together, these findings underscore the value of time-sensitive, within-person monitoring for understanding the dynamic mechanisms through which suicidality emerges and for advancing more personalized approaches to suicide risk detection and intervention.

## Supporting information

Supplementary Material SI DTW paper

## Data Availability

The data used in this study were obtained from the NESDA, NESDO, and NOCDA cohorts. Due to privacy restrictions and ethical regulations, the raw data are not publicly available. Access to the data can be requested through the respective study secretariats, subject to approval by their scientific committees and in accordance with their data access protocols.

## References

1. Lim, K.S., et al., Global Lifetime and 12-Month Prevalence of Suicidal Behavior, Deliberate Self-Harm and Non-Suicidal Self-Injury in Children and Adolescents between 1989 and 2018: A Meta-Analysis. Int J Environ Res Public Health, 2019. 16(22).

2. Fried, E.I. and R.M. Nesse, Depression is not a consistent syndrome: An investigation of unique symptom patterns in the STAR*D study. J Affect Disord, 2015. 172: p. 96–102.

3. Franklin, J.C., et al., Risk factors for suicidal thoughts and behaviors: A meta-analysis of 50 years of research. Psychol Bull, 2017. 143(2): p. 187–232.

4. Oquendo, M.A., et al., Lifetime Suicide Attempts in Otherwise Psychiatrically Healthy Individuals. JAMA Psychiatry, 2024. 81(6): p. 572–578.

5. Turecki, G. and D.A. Brent, Suicide and suicidal behaviour. Lancet, 2016. 387(10024): p. 1227–39.

6. O’Connor, R.C. and O.J. Kirtley, The integrated motivational-volitional model of suicidal behaviour. Philos Trans R Soc Lond B Biol Sci, 2018. 373(1754).

7. de Beurs, D., et al., Symptoms of a feather flock together? An exploratory secondary dynamic time warp analysis of 11 single case time series of suicidal ideation and related symptoms. Behav Res Ther, 2024. 178: p. 104572.

8. de Beurs, D., et al., A network perspective on suicidal behavior: Understanding suicidality as a complex system. Suicide Life Threat Behav, 2021. 51(1): p. 115–126.

9. Raifman, J., et al., Economic precarity, loneliness, and suicidal ideation during the COVID-19 pandemic. PLoS One, 2022. 17(11): p. e0275973.

10. O’Connor, R.C., et al., Mental health and well-being during the COVID-19 pandemic: longitudinal analyses of adults in the UK COVID-19 Mental Health & Wellbeing study. Br J Psychiatry, 2021. 218(6): p. 326–333.

11. Efstathiou, V., et al., A one-year longitudinal study on suicidal ideation, depression and anxiety during the COVID-19 pandemic. Asian J Psychiatr, 2022. 73: p. 103175.

12. Brody, L.R., & Hall, J. A., Gender and emotion in context. Handbook of emotions, 2008. 3: p. 395–408.

13. Ha, H. and E.J. Shim, Do the Relative Importance and Pattern of Correlates of Suicidal Ideation Vary by Age and Gender? Network Analyses. Int J Psychol, 2025. 60(3): p. e70049.

14. Holman, M.S. and M.N. Williams, Suicide Risk and Protective Factors: A Network Approach. Arch Suicide Res, 2022. 26(1): p. 137–154.

15. Kopland, M.C.G. and E.J. Giltay, Dynamic Time Warp (DTW) as a scalable, data-efficient, and clinically relevant analysis of dynamic processes in patients with psychiatric disorders: a tutorial. J Eat Disord, 2025. 13(1): p. 230.

16. Molenaar, P.C.M., & Campbell, C. G., The New Person-Specific Paradigm in Psychology. . Current Directions in Psychological Science, 2009. 18(2): p. 112–117.

17. McNally, R.J., Network Analysis of Psychopathology: Controversies and Challenges. Annu Rev Clin Psychol, 2021. 17: p. 31–53.

18. Borsboom, D., et al., Network analysis of multivariate data in psychological science. Nature Reviews Methods Primers, 2021. 1(1).

19. van den Brink, B., et al., Experience sampling of suicidality, religiosity and spirituality in depression: Network analyses using dynamic time warping. J Affect Disord, 2024. 360: p. 354–363.

20. Hallensleben, N., et al., Predicting suicidal ideation by interpersonal variables, hopelessness and depression in real-time. An ecological momentary assessment study in psychiatric inpatients with depression. Eur Psychiatry, 2019. 56: p. 43–50.

21. Kleiman, E.M., et al., Examination of real-time fluctuations in suicidal ideation and its risk factors: Results from two ecological momentary assessment studies. J Abnorm Psychol, 2017. 126(6): p. 726–738.

22. Penninx, B., et al., Cohort profile of the longitudinal Netherlands Study of Depression and Anxiety (NESDA) on etiology, course and consequences of depressive and anxiety disorders. J Affect Disord, 2021. 287: p. 69–77.

23. Comijs, H.C., et al., The Netherlands study of depression in older persons (NESDO); a prospective cohort study. BMC Res Notes, 2011. 4: p. 524.

24. Schuurmans, J., et al., The Netherlands Obsessive Compulsive Disorder Association (NOCDA) study: design and rationale of a longitudinal naturalistic study of the course of OCD and clinical characteristics of the sample at baseline. Int J Methods Psychiatr Res, 2012. 21(4): p. 273–85.

25. Rush, A.J., et al., The 16-Item Quick Inventory of Depressive Symptomatology (QIDS), clinician rating (QIDS-C), and self-report (QIDS-SR): a psychometric evaluation in patients with chronic major depression. Biol Psychiatry, 2003. 54(5): p. 573–83.

26. Beck, A.T., et al., An inventory for measuring clinical anxiety: psychometric properties. J Consult Clin Psychol, 1988. 56(6): p. 893–7.

27. De Jong Gierveld, J. and T. Van Tilburg, The De Jong Gierveld short scales for emotional and social loneliness: tested on data from 7 countries in the UN generations and gender surveys. Eur J Ageing, 2010. 7(2): p. 121–130.

28. Meyer, T.J., et al., Development and validation of the Penn State Worry Questionnaire. Behav Res Ther, 1990. 28(6): p. 487–95.

29. Veenhoven, R., Ehrhardt, J., The cross-national pattern of happiness: Test of predictions implied in three theories of happiness. Soc Indic Res 1995. 34: p. 33–68.

30. Pan, K.Y., et al., The mental health impact of the COVID-19 pandemic on people with and without depressive, anxiety, or obsessive-compulsive disorders: a longitudinal study of three Dutch case-control cohorts. Lancet Psychiatry, 2021. 8(2): p. 121–129.

31. van der Does, F., et al., Dynamic time warp versus vector autoregression models for network analyses of psychological processes. Sci Rep, 2025. 15(1): p. 11720.

32. Pavulans, K.S., et al., Being in want of control: Experiences of being on the road to, and making, a suicide attempt. Int J Qual Stud Health Well-being, 2012. 7.

33. Aschenbrenner, L.M., et al., Exploring suicidal behaviour through implicit identity and control biases: Findings from the Death-Implicit Association Test and its novel control-adaptation. Compr Psychiatry, 2025. 142: p. 152621.

34. Forkmann, T. and T. Teismann, Entrapment, perceived burdensomeness and thwarted belongingness as predictors of suicide ideation. Psychiatry Res, 2017. 257: p. 84–86.

35. O’Connor, R.C. and M.K. Nock, The psychology of suicidal behaviour. Lancet Psychiatry, 2014. 1(1): p. 73–85.

36. Motillon-Toudic, C., et al., Social isolation and suicide risk: Literature review and perspectives. Eur Psychiatry, 2022. 65(1): p. e65.

37. De Leo, D., Late-life suicide in an aging world. Nat Aging, 2022. 2(1): p. 7–12.

38. Michaels, M.S., et al., Total sleep time as a predictor of suicidal behaviour. J Sleep Res, 2017. 26(6): p. 732–738.

39. Rogers, M.L., J.Y. Gorday, and T.E. Joiner, Examination of characteristics of ruminative thinking as unique predictors of suicide-related outcomes. J Psychiatr Res, 2021. 139: p. 1–7.

40. Bentley, K.H., et al., Anxiety and its disorders as risk factors for suicidal thoughts and behaviors: A meta-analytic review. Clin Psychol Rev, 2016. 43: p. 30–46.

41. Moreno-Muñoz, P., Romero-Medrano, L., Moreno, Á., Herrera-López, J., Baca-García, E., & Artés-Rodríguez, A., Passive detection of behavioral shifts for suicide attempt prevention. . arXiv preprint arXiv:2011.09848., 2020.

42. Burger, J., et al., A clinical PREMISE for personalized models: Toward a formal integration of case formulations and statistical networks. J Psychopathol Clin Sci, 2022. 131(8): p. 906–916.

43. Rothwell, P.M., External validity of randomised controlled trials: “to whom do the results of this trial apply?”. Lancet, 2005. 365(9453): p. 82–93.

44. Granger, C.W.J., Investigating Causal Relations by Econometric Models and Cross-spectral Methods. Econometrica, 1969. 37(3): p. 424–438.

45. Molenaar, P.C.M., A Manifesto on Psychology as Idiographic Science: Bringing the Person Back Into Scientific Psychology, This Time Forever. . Measurement: Interdisciplinary Research and Perspectives, 2004. 2(4): p. 201–218.

