## Supplementary Material SI DTW paper for "Within-person temporal alignment shows symptom co-fluctuations and early precursors of suicidal ideation"

This appendix has been provided by the authors to give readers additional information about their work.

**Supplementary materials**

Manuscript title: Within-person temporal alignment shows symptom co-fluctuations and early precursors of suicidal ideation.

Authors: A.J.C. van der Slot, C. Boonmann, A.A.L. Kok, M.W.M. Gijzen, M. Eikelenboom, D. de Beurs, B.W.J.H. Penninx and E.J. Giltay

*Overview of supplementary content:*	page

Figure 1	Flow chart of inclusion/exclusion process	iii

Table 1	Baseline characteristics of included vs. excluded participants	iv-v

*DTW sample script*	vi

*COVID-19*	COVID-19-specific exposures and responses (21 items)	vii

Figure 2	Dynamic alignment of depressive symptoms with SI across sex and age groups	vii

Figure 3	Dynamic alignment of anxiety symptoms with SI across sex and age groups	ix

Figure 4	Dynamic alignment of loneliness symptoms with SI across sex and age groups. x

Figure 5	Dynamic alignment of worry symptoms with SI across sex and age groups.	xi

Figure 6	Dynamic alignment of COVID-19 specific stressors with SI across sex and	xii
age groups

**Supplementary Figure 1**: Flow chart of inclusion/exclusion process.

2981 NESDA participants were assessed for eligibility

510 NESDO participants were assessed for eligibility

29.7% (1162/3910) were excluded due to no consent given for follow-up

2748 participants were invited during COVID-19 pandemic

59.1% (1623/2748) were unable to complete at least one COVID-19 questionaire

1125 participants eligible for suicidal ideation (SI) change analysis

72.7% (818/1125) were excluded for not showing any change in SI

307 were used for the current study

419 NOCDA participants were assessed for eligibility

**Table 1.** Baseline characteristics of included vs. excluded participants from the NESDA/NESDO/NOCDA cohorts.

|  | Current study | | P** |
| --- | --- | --- | --- |
|  | Included (n=307) | Excluded * (n=818) |  |
| **Socio-demographics** |  |  |  |
| Age (SD) | 44.8 (12.1) | 44.6 (13.2) | .735 |
| Female sex (%) | 189 (61.6%) | 535 (65.4%) | .130 |
| High level of education (%) | 118 (38.4%) | 372 (45.5%) | .063 |
| **Comorbidity** |  |  |  |
| Lifetime disorder (%) | 307 (100%) | 552 (67.5%) | <.001 |
| Number of lifetime disorders (SD) | 3.38 (1.38) | 1.76 (1.67) | <.001 |
| Chronicity (SD) | 2.33 (0.77) | 1.24 (1.08) | <.001 |
| **Vaccination attitude** |  |  |  |
| Willing to vaccinate | 220 (87.0%) | 585 (87.7%) | .825 |
| **Functional scores** |  |  |  |
| Avg QIDS pre COV (IQR) | 8.8 (5.8-11.5) | 3.3 (2.0-6.0) | <.001 |
| Avg BAI pre COV (IQR) | 11.6 (6.1-17.1) | 4.0 (1.6-8.0) | <.001 |
| Avg PSWQ pre COV (IQR) | 33.8 (26.3-41.4) | 20.8 (15.0-29.2) | .284 |
| Avg JGLS pre COV (IQR) | 3.0 (1.0-5.0) | 1 (0.0-2.5) | <.001 |
| Avg QIDS during COV (IQR) | 8.7 (6.0-13.0) | 3.0 (2.0-5.0) | <.001 |
| Avg BAI during COV (IQR) | 11.7 (5.4-19.7) | 2.3 (0.0-6.4) | <.001 |
| Avg PWQR during COV (IQR) | 35.0 (27.7-43.3) | 21.0 (13.7-28.3) | .157 |
| Avg JGLS during COV (IQR) | 4.0 (2.3-5.6) | 1.3 (0.7-2.7) | <.001 |

*Participants without a lifetime psychiatric diagnosis and those without any change in suicidal ideation (IDS item 12) during the pandemic were excluded from the analytical sample. **p was estimated by the Chi square test for categorical variables, t-tests for normally distributed variables and the Kruskal-Wallis test for non-normally distributed variables. Comorbidity is assessed through CIDI (Composite International Diagnostic Interview*)* diagnostic interviews based on DSM-VI criteria. IQR denotes Interquartile range (25^th^ and 75^th^ percentiles); QIDS denotes quick inventory of depressive symptomatology; BAI denotes Beck Anxiety Inventory; PWQR denotes Penn State Worry Questionnaire; JGLS denotes de Jong-Gierveld Loneliness Scale. Range: QIDS (0-27), BAI (0-63), PWQR (16-90), JGLS (0-6).

**Sample R script** (<https://osf.io/gbaw2/>)

Note on Script and Methodology: > This sample script accompanied an earlier DTW paper we published. Although it reflects different outcomes, it utilizes the same mathematical principle of non-linear temporal alignment applied in this study. For a comprehensive walkthrough of the DTW methodology, parameter selection (such as the Sakoe–Chiba band), and clinical interpretation, we refer to the following tutorial:

Kopland MCG, Giltay EJ. Dynamic Time Warp (DTW) as a scalable, data-efficient, and clinically relevant analysis of dynamic processes in patients with psychiatric disorders: a tutorial. J Eat Disord. 2025;13(1):230.

**COVID-19-specific exposures and responses (English Translation)**

Item (response options: 1=totally disagree – 5=totally agree)

Perceived Impact (mental burden – 9 items)

- Because of this period the quality of my sleep is worse
- This period makes me consume more snacks and sweets
- This period makes me drink more alcohol
- This period makes me more emotional
- In this period I’m having more nightmares
- In this period it’s hard to concentrate
- In this period I’m more often lonely
- This period makes me sad
- In this period I’m more often irritable and tense

Fear of Infection – 6 items

- I fear to become infected with corona
- Because of the threat of the virus I dont leave my home anymore
- This period makes me fearful
- I intensively follow the news about the virus through TV newspaper and or social media
- Because of the threat of the virus I am anxious of getting close to other people
- I strictly follow the rules to prevent contamination and spread of the virus

Positive Coping – 5 items

- In this period I feel more connected to society
- It is no problem to enjoy myself while being at home more often
- I have confidence that the Netherlands will overcome this crisis
- Despite the virus I stay active (household tasks, gardening, walking, sporting, yoga)
- Despite the virus I actively maintain (via phone or online) contacts with friends

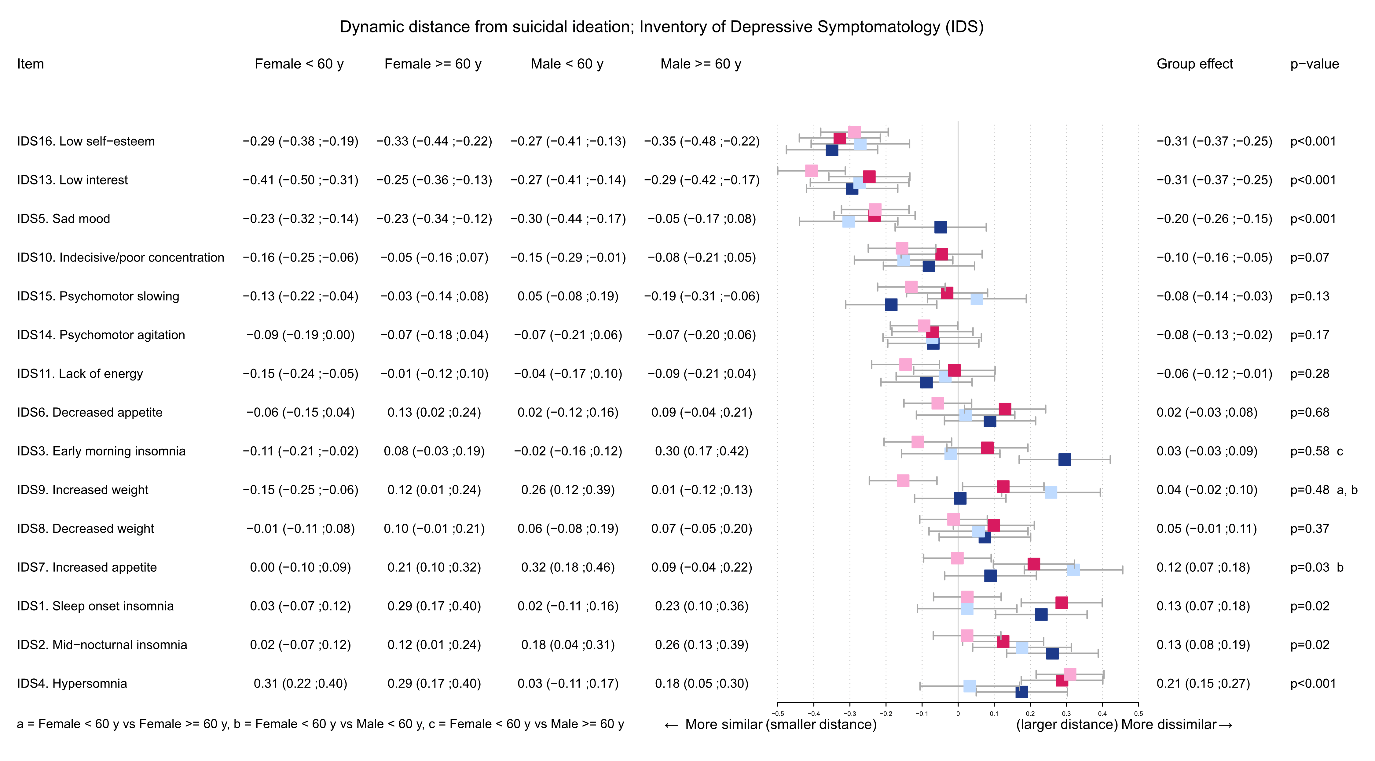
**Supplementary Figure 3.** Dynamic alignment of depressive symptoms with SI across sex and age groups.

Undirected DTW analysis was used to compute the temporal distance between depressive symptoms (IDS items) and SI. Group effects by sex (male, female) and age (<65 years, ≥65 years) are indicated. Smaller distances indicate greater co-fluctuation over time. P-values for the item compared with SI are displayed to the right of each item, with p < 0.05 considered statistically significant. Group effects are provided and significance is indicated with a letter on the right of the overall p value of the item. Items low energy (IDS5), low self-esteem (IDS16), and sad mood (IDS13) showed the closest temporal association with SI across groups (p<0.001). In contrast, somatic symptoms such as hypersomnia (IDS4), *mid-nocturnal insomnia* (IDS2) sleep onset insomnia (IDS1), and increased appetite (IDS7) were least aligned with SI. Significant group differences were observed for Increased appetite (IDS7), Increased weight (IDS9) and *early morning insomnia* (IDS3).

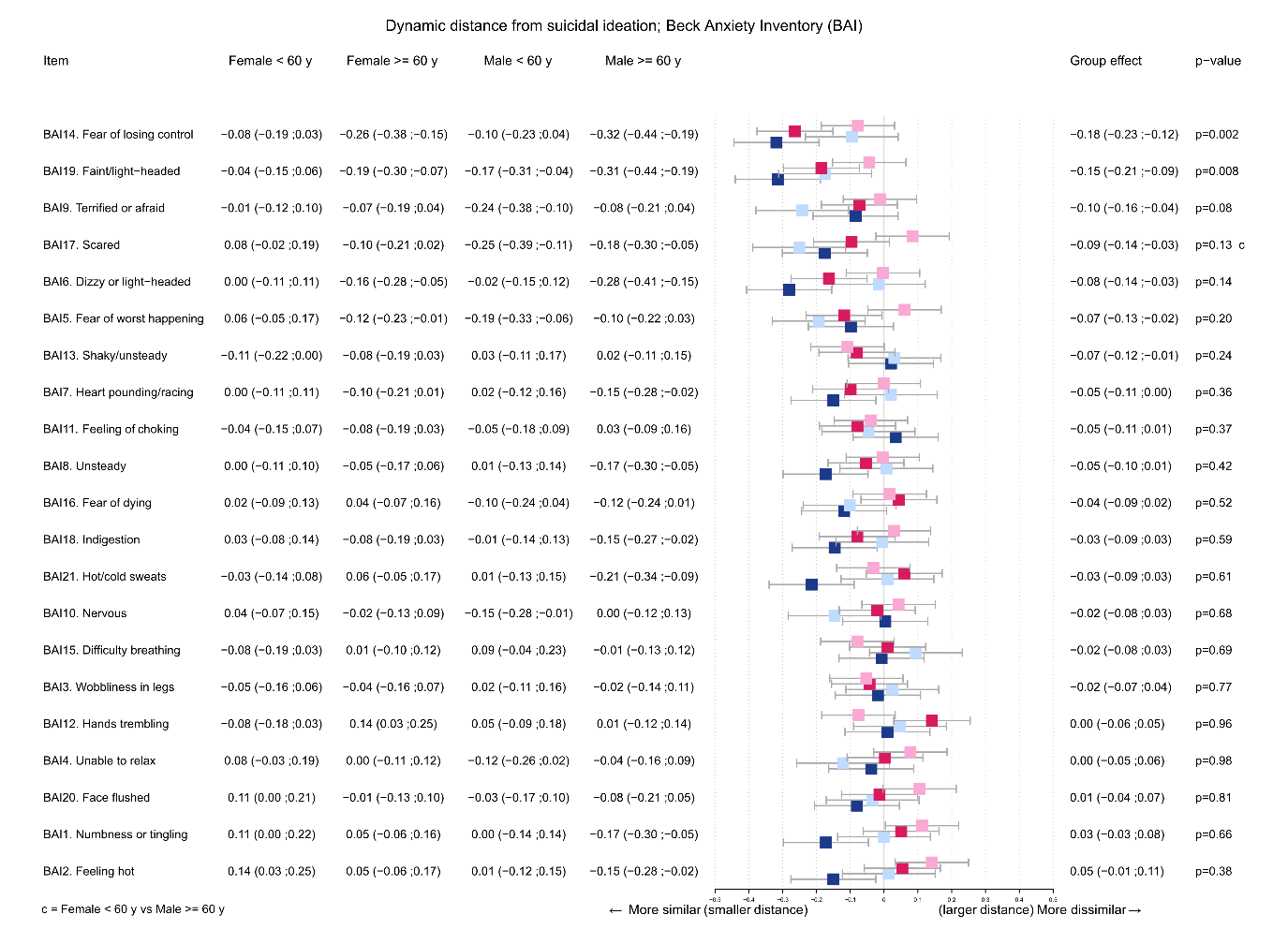
**Supplementary Figure 4.** Dynamic alignment of anxiety symptoms with SI across sex and age groups.
Undirected DTW analysis was used to compute the temporal distance between anxiety symptoms (BAI items) and SI. Smaller distances indicate greater co-fluctuation over time. P-values for the item compared with SI are displayed to the right of each item, with p < 0.05 considered statistically significant. Group effects are provided and significance is indicated with a letter on the right of the overall p value of the item. The cognitive-affective symptom fear of losing control (BAI14, p = 0.005) and faint/light-headedness (BAI19, p = 0.007) were significantly aligned with SI. A significant group effect was found for scared (BAI17), reflecting differences between adult males (65-) and adult females (65-). No other significant subgroup differences were found.

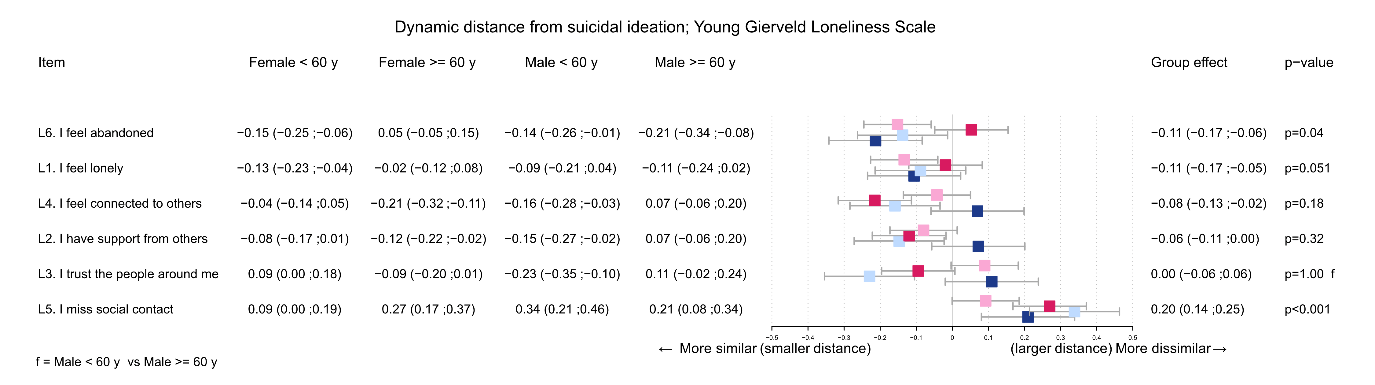
**Supplementary Figure 5.** Dynamic alignment of loneliness symptoms with SI across sex and age groups.
Dynamic time warping was applied to loneliness items from the De Jong Gierveld Loneliness Scale to examine their alignment with SI. Smaller distances indicate greater co-fluctuation over time. P-values for the item compared with SI are displayed to the right of each item, with p < 0.05 considered statistically significant. Group effects are provided and significance is indicated with a letter on the right of the overall p value of the item. Emotional loneliness item feeling abandoned was significantly aligned with SI and feeling lonely was moderately aligned with SI p = 0.051, but no significant group differences were observed (all p > 0.05). Missing social contacts demonstrated the least similarity from SI.

**Supplementary Figure 6.** Dynamic alignment of worry symptoms with SI across sex and age groups.
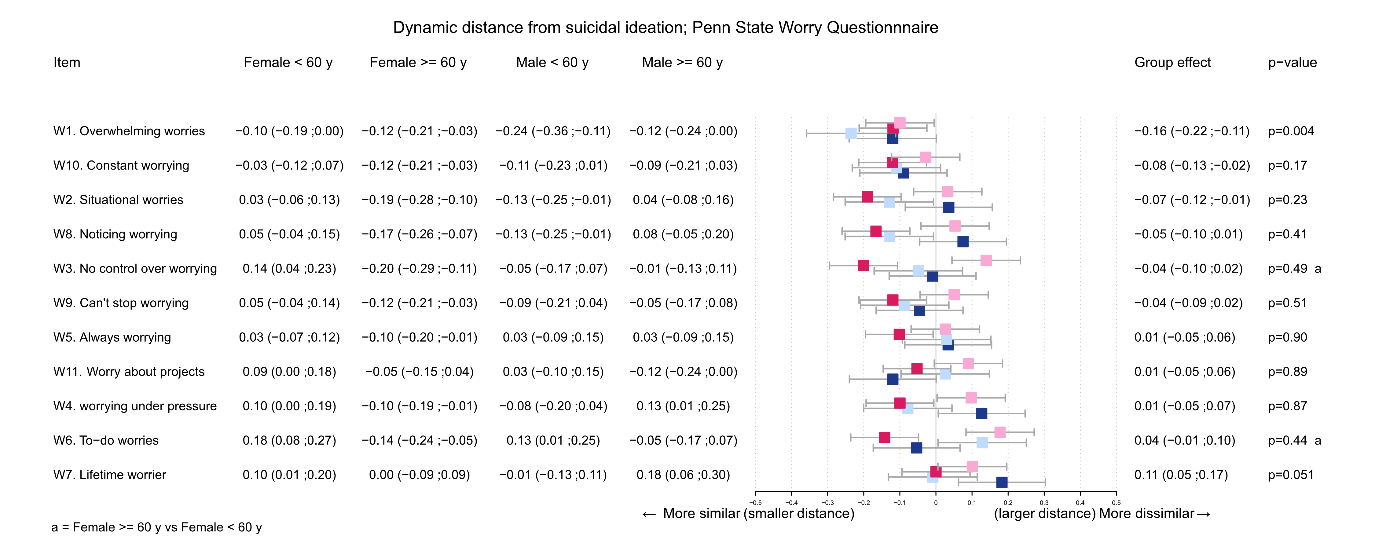

Undirected dynamic time warping (DTW) analysis was used to compute the temporal distance between individual items of the Penn State Worry Questionnaire (PSWQ) and SI. Smaller distances indicate greater co-fluctuation over time. P-values for the item compared with SI are displayed to the right of each item, with p < 0.05 considered statistically significant. Group effects are provided and significance is indicated with a letter on the right of the overall p value of the item. Among all items, only "overwhelming worries" (W1) demonstrated a significant association with SI (p = 0.004), with smaller distances indicating closer temporal alignment. Group effects were observed for "no control over worrying" (W3; p = 0.049) and "overwhelming worries" (W1; p = 0.004), showing greater alignment with SI in younger men (<60 years) compared to women.

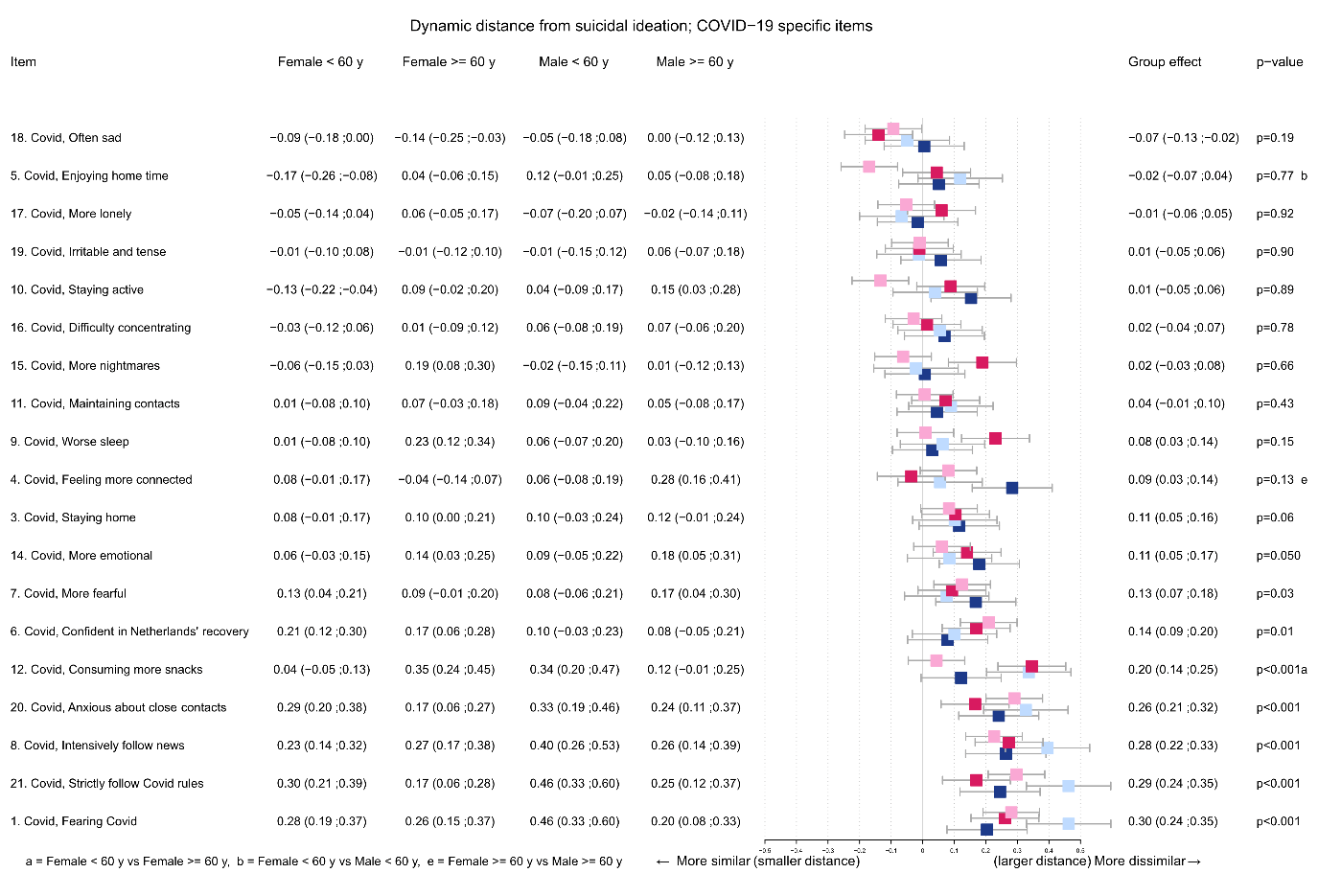
**Supplementary Figure 7.** Dynamic alignment of COVID-19 specific stressors with SI across sex and age groups.
Undirected DTW analysis was used to compute the temporal distance between COVID-19-related items and SI. Smaller distances indicate stronger temporal alignment. P-values indicate group-level similarity between each item and SI, with p < 0.05 considered statistically significant. Group differences by age and sex are shown on the right and are denoted with lettered superscripts (see legend below). No COVID-19 items were significantly aligned with SI (all p > 0.05), suggesting low temporal association overall. Significant group effects were found for four items: fearing COVID-19 (Item 1, group f), strictly following COVID rules (Item 21, p < 0.001, group d), intensively following the news (Item 8, p < 0.001, group e), and anxiety about close contacts (Item 20, p < 0.001, group e).
